# Integrating Infection Burden and Multimodal Biomarkers for Early Detection of Alzheimers Disease: A Sheaf-ML Framework

**DOI:** 10.1101/2025.11.10.25339915

**Authors:** Lokendra S. Thakur, Gurpreet Bharj, Duc Thanh Nguyen, Manish Saroya, Bushra Malik

## Abstract

Alzheimers disease (AD) remains a major global health challenge, with growing evidence linking chronic infections, immune aging, and neurodegeneration. Grounded in the *Antimicrobial Protection Hypothesis*, this study introduces a sheaf-theoretic machine learning framework, **Sheaf-ML**, for integrating multimodal health data and assessing infection-related cognitive risk. Sheaf-ML constructs a **unified patient-level representation** that coherently combines diverse data streamsincluding serological infection markers, cognitive assessments, cardiovascular and metabolic measures, nutritional and behavioral evaluationswhile preserving the intrinsic structure and relationships of each modality. Applying this framework to the **Harmonized LASI-DAD** dataset (*N* = 6168), we modeled six clinically motivated domains (Infection, Cognition, Mental Health, Cardiovascular, Nutrition, and Demographics) and integrated them into a topologically consistent representation using learnable cross-domain mappings and consistency constraints. The sheaf-integrated embeddings revealed clinically meaningful interactions: infection burden was linked with cardiovascular, nutritional, and cognitive outcomes, highlighting system-level coordination across modalities. Using these embeddings, Sheaf-ML produced interpretable patient-level predictions and identified the most influential features both globally and individually. We further derived an **Infection Burden Index (IBI)**, which quantified patient-level infection-related risk. Patients exceeding the 80^th^ percentile were flagged as early-warning cases, corresponding to approximately 20% of the cohort, demonstrating actionable stratification for clinical monitoring. This study provides the first empirical evidence that sheaf-based architectures can integrate multimodal health data in a clinically interpretable manner, uncover biologically meaningful interactions, and support patient-specific risk prediction. By linking population-level patterns with individualized insights, Sheaf-ML establishes a foundation for scalable, interpretable, and equitable precision models of infection-related cognitive decline in Alzheimers disease.

## 1 Introduction

Alzheimers Disease (AD) represents a critical global health challenge, with current diagnostic tools often failing to capture the early transition from Mild Cognitive Impairment (MCI) to AD.^**1**^ Increasing evidence implicates chronic infectionsparticularly herpes simplex virus type 1 (HSV-1) and cytomegalovirus (CMV)as contributors to neurodegenerative progression through immune-inflammatory pathways, including NLRP3 inflammasome activation, microglial dysregulation, and amyloid-beta (A^*β*^) aggregation.^**2**^ These findings emphasize the need for integrative, mechanistically informed models capable of identifying infection-related risk factors prior to irreversible cognitive decline.

To address this gap, we introduce a **Sheaf-Theoretic Infection-Tracking Framework** for multimodal data integration and cognitive risk modeling. **Sheaf theory** provides a principled topological and categorical foundation for coherently fusing locally defined, heterogeneous data sources,^**3**^ while **topos theory** enables logical reasoning and constraint enforcement across diverse data modalities.^**4, 5**^ Building on ideas from the **Seifert–van Kampen theorem**^**6, 7**^and **fibered categories**,^**8, 9**^ our framework preserves structural relationships across patient-specific data histories, enabling both interpretability and generalization.

Within this formalism, we define a **fibered multimodal data space** in which each patient constitutes a fiber populated by modality-specific sheaves (e.g., Infection, Cognition, Nutrition, Cardiovascular, Mental Health). Learnable restriction maps between these domains capture cross-modality transformations, while a gluing-consistency loss enforces coherence across overlapping data subsets. Operating within this space, the **Sheaf Machine Learning (Sheaf-ML)** architecture integrates heterogeneous, asynchronous, and incomplete data into a unified, interpretable representation.

Our **results on the Harmonized LASI-DAD dataset** demonstrate the feasibility, effectiveness, and clinical interpretability of the sheaf-based framework. By integrating six biologically grounded modalitiesdemographics, cognition, mental health, cardiovascular function, nutrition, and infectioninto a unified 111-dimensional representation, **Sheaf-ML** effectively captures both intra-domain and cross-domain interactions. The model identified cognitive vulnerability and infection-related risk, while maintaining interpretability through global feature contributions, patient fiber visualizations, and neighborhood-level embeddings. In particular, the **Infection Burden Index (IBI)** highlighted biologically meaningful links among infection, cardiovascular, and cognitive domains, supporting early-warning identification for high-risk individuals. These results validate the applicability of sheaf-based architectures for multimodal health data integration and clinically interpretable outcome prediction in large, real-world populations.

Building on this foundation, we define the **Infection Burden Index (IBI)**a dynamic, interpretable biomarker derived from integrated serological, physiological, and cognitive featuresthat captures infection-related neurocognitive risk. The IBI is envisioned as a scalable tool for early detection, individualized intervention, and population-specific risk stratification in AD, forming the basis for future clinical validation studies.

## 2 Objective

The primary objective of this study is to establish and validate a **multimodal, mathematically grounded framework** that leverages **Sheaf Machine Learning (Sheaf-ML)** to integrate infection-related, physiological, behavioral, and neurocognitive data into a unified representational space. Within this **fibered multimodal data space**, each modality (e.g., Infection, Cognition, ADL, Nutrition, Cardiovascular, Mental Health) contributes locally coherent information that is globally synthesized through learnable restriction maps and gluing-consistency constraints. From this integrated structure, we aim to derive the **Infection Burden Index (IBI)**a dynamic, interpretable biomarker quantifying infection-related neurocognitive risk. The IBI is envisioned as a predictive tool and clinical decision-support metric to guide personalized interventions that may delay or mitigate the progression from Mild Cognitive Impairment (MCI) to Alzheimers disease (AD).

## 3 Roadmap to Accomplish the Objective

This study follows a three-phase roadmap toward developing a mathematically principled, infection-aware multimodal tracking framework for Alzheimers disease. In **Phase I**, we develop the **Sheaf-ML computational framework** based on sheaf theory and fibered-category formalisms to integrate heterogeneous biomedical modalities. The model operationalizes learnable restriction maps and gluing losses to capture structural dependencies across modalities. Preliminary implementation on the **Harmonized LASI-DAD dataset**comprising seven biologically grounded modalities and 147 featuresdemonstrates that sheafintegrated representations improve cognitive impairment classification accuracy and yield interpretable topological structures that reflect inter-domain relationships. In **Phase II**, we formalize the **Infection Burden Index (IBI)** as a sheaf-derived multimodal biomarker capturing infection-associated cognitive vulnerability. The IBI will be validated across multi-cohort datasets (ADNI, OASIS, NACC, ICMR, AMP-AD), assessing reproducibility, robustness to data missingness, and cross-population generalizability. Logic-regularized losses encoding biomedical priors (e.g., infection burdencognition linkages) will enhance interpretability and clinical plausibility. Finally, **Phase III** outlines a translational step toward clinical implementation, involving the design of a pilot randomized study to evaluate the feasibility and clinical utility of IBI-guided personalized interventions in Indian cohorts (NIMHANS, LASI). This phase will inform scalability, cost-effectiveness, and integration within real-world dementia care pipelines.

Together, these phases delineate a rigorous translational trajectory from topological data modeling to actionable clinical applications, establishing the Infection Burden Index (IBI) as a scalable, interpretable, and population-adaptive biomarker for infection-modulated neurodegeneration in Alzheimers disease.

## 4 Significance

Alzheimers Disease (AD) remains one of the most pressing challenges in clinical neuroscience, with limited strategies for early detection and precision intervention. Current diagnostic paradigms rely primarily on imaging and cognitive testing, which often detect disease only after substantial neurodegeneration has occurred. Concurrently, growing evidence implicates latent infectionsparticularly herpesviruses such as HSV-1 and CMVas potential contributors to AD pathogenesis through mechanisms involving chronic neuroinflammation, NLRP3 inflammasome activation, and amyloid-beta (A*β*) aggregation. Despite these insights, there is currently no validated computational or clinical framework capable of quantifying infection-related neurocognitive risk in a dynamic, individualized manner. This project addresses these critical gaps through the development of a **Sheaf-Theoretic Machine Learning (Sheaf-ML)** framework that mathematically integrates multimodal health data within a **fibered multimodal data space**. Using the principles of sheaf theory, topos logic, and fibered categories, this framework enables coherent fusion of heterogeneous modalitiesincluding serological infection markers, cardiovascular and metabolic measures, behavioral and cognitive assessments, and wearable sensor datawhile preserving modality-specific structure and relationships. Preliminary implementation on the **Harmonized LASI-DAD dataset** demonstrates that Sheaf-ML yields improved classification performance for cognitive impairment, interpretable topological representations, and biologically meaningful embeddings that reveal inter-domain dependencies (e.g., infectioncognition links). Building on these results, we introduce the **Infection Burden Index (IBI)**a sheaf-integrated, interpretable biomarker that dynamically quantifies infection-associated neurocognitive risk. By providing a unified and mathematically consistent measure of multimodal health burden, the IBI represents a paradigm shift from late-stage reactive diagnosis to early-stage, mechanism-informed intervention. Clinically, this framework will enable the identification of individuals at heightened risk for cognitive decline, offering actionable insights for personalized care pathways and preventive strategies.

Beyond Alzheimers Disease, the Sheaf-ML infrastructure establishes a generalizable and explainable mathematical foundation for multimodal data integration in chronic neurodegenerative and immune-mediated conditions. Its emphasis on topological structure, interpretability, and population adaptability positions it as a scalable precision health platform for data-driven, context-aware decision support in translational medicine.

## 5 Innovation

This study introduces a transformative framework that unites advanced mathematical modeling, mechanistic neuroscience, and translational data science to address a critical gap in early Alzheimers Disease (AD) risk assessment. Its innovation spans three complementary dimensions: **1. Mathematical Innovation:** We present a fundamentally new paradigm for biomedical data integration grounded in **sheaf theory, fibered categories**, and **topos-theoretic reasoning**. These algebraic-topological tools enable principled fusion of multimodal, incomplete, and asynchronous health data while preserving their local structural and contextual relationships. Through learnable *restriction maps* and *gluing-consistency constraints*, the **Sheaf Machine Learning (Sheaf-ML)** framework generalizes beyond traditional early-or late-fusion models, which typically discard modality interdependencies through vectorization. The resulting **fibered multimodal data space** provides a mathematically coherent foundation for interpretable, noise-resilient learning across diverse biomedical modalities. **2. Conceptual Innovation:** We define the **Infection Burden Index (IBI)** as a new class of *topologically informed, sheaf-derived biomarkers*. Rather than treating infections as isolated covariates, the IBI models them as dynamic, interacting processes contributing to neurocognitive decline through immune and metabolic pathways. This sheaf-integrated index captures the evolving interplay between infection, inflammation, and cognition within a unified representational geometry. The approach moves beyond correlational modeling toward mechanistically informed, logically constrained inferencelinking biological theory with machine-learned structure. **3. Clinical Innovation:** This work represents the first application and planned clinical translation of a **sheaf-based, multimodal infection-tracking framework** for cognitive health monitoring. Preliminary validation on the **Harmonized LASI-DAD dataset** demonstrates both predictive accuracy and structural interpretability, providing a foundation for real-world deployment. Through integration with wearable biosensors, mobile cognitive assessments, and clinical data streams, the system enables continuous, patient-centered tracking of infection-related neurocognitive risk. Designed for interoperability and transparency, the Sheaf-ML platform supports **real-time decision support**, advancing equitable and personalized dementia care.

Together, these innovations establish a novel scientific and clinical framework for understanding, detecting, and managing infection-modulated neurodegeneration. By merging rigorous mathematical abstraction with translational applicability, this project defines a new class of interpretable, topologically structured models for precision medicine in Alzheimers Disease and beyond.

## 6 Data Sources and Availability

Multimodal datasets have been verified from Indian and U.S. aging cohorts (Table 1), encompassing cognitive, serological, neuroimaging, and wearable modalities. These include LASI and ICMR for Indian populations, and ADNI, NACC, AMP-AD, and MIMIC-IV among global repositories, collectively enabling rigorous cross-cohort validation of the Infection Burden Index (IBI).

**Table 1.**
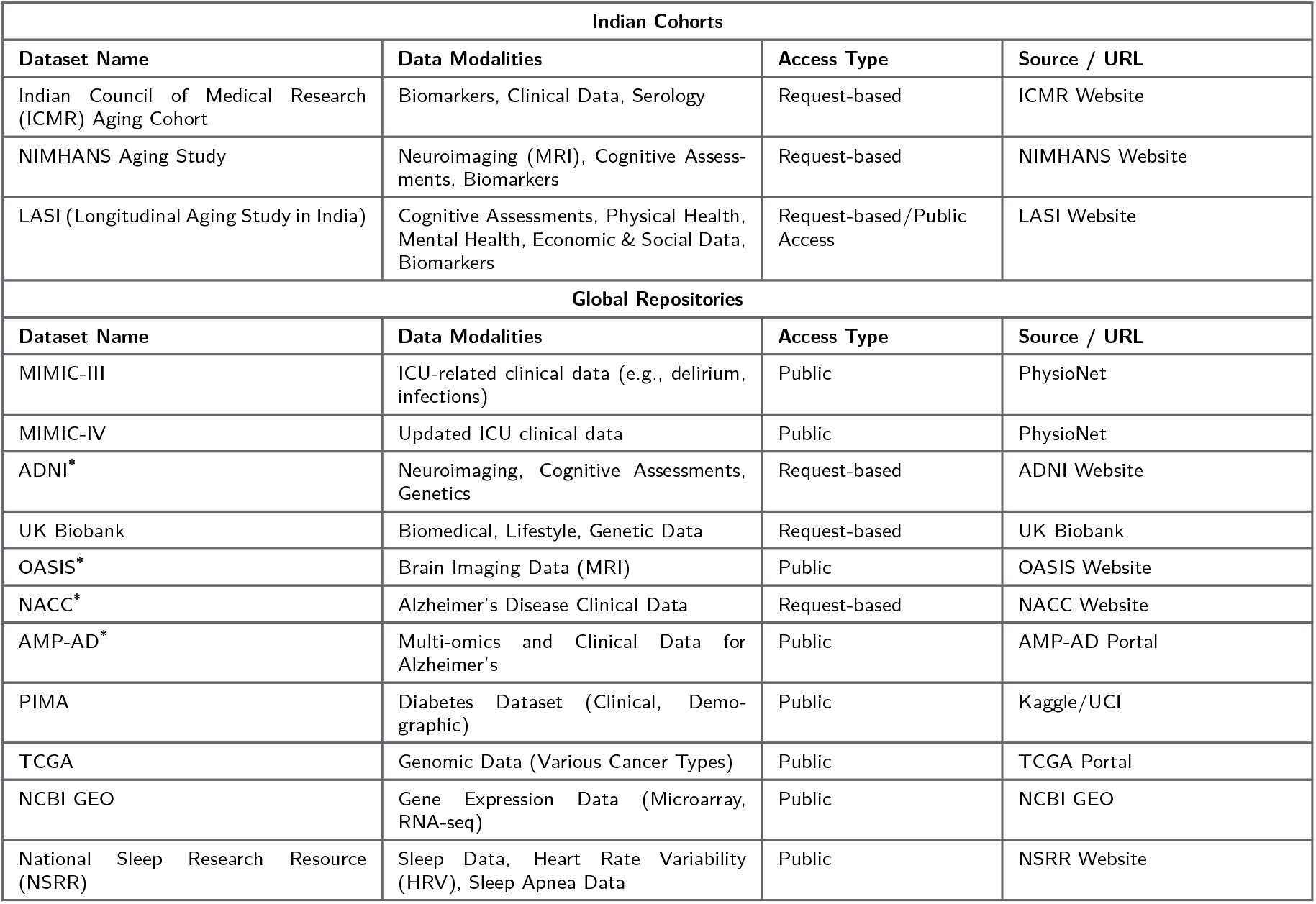
Data Modalities for Indian and Global Cohorts.

**Table 2.**
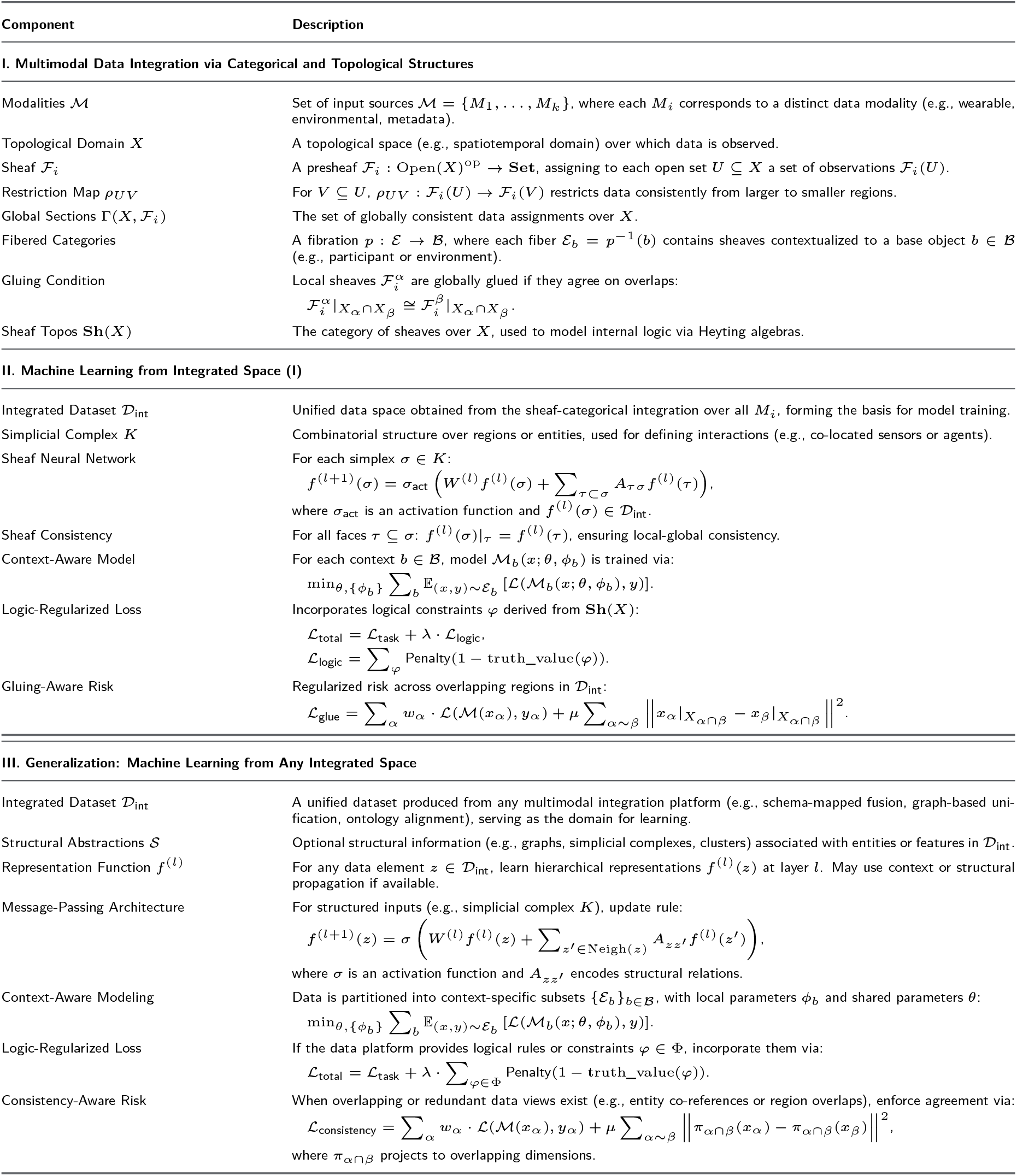
Mathematical formulation of the framework. The machine learning model is trained on the sheaf-integrated data space 𝒟_int_ or any other multimodal integration platform, incorporating logical and topological constraints from categorical structures.

### 6.1 Data Description

This analysis uses data or information from the Harmonized LASI-DAD programming codes and Codebook, Version B.1 as of Feburary 2025, developed by the Gateway to Global Aging Data (DOI: https://doi.org/10.25553/h5wx-ay45). The development of the Harmonized LASI-DAD was funded by the National Institute on Aging (Wave 1: R01 AG0330153, RF1 AG055273, U01 AG064948 Wave 2: R01 AG051125, U01 AG065958, R01 AG030153). For more information about the Harmonization project, please refer to “https://g2aging.org/

The Harmonized LASI-DAD dataset^**10**^ provides detailed cognitive assessments, informant interviews, and health measures that have been harmonized with other international studies in the Health and Retirement Study (HRS) family, such as the U.S. HRS-DAD and the English Longitudinal Study of Ageing (ELSA-DAD). The purpose of LASI-DAD is to estimate the prevalence, determinants, and impact of cognitive impairment and dementia in India and to facilitate cross-national comparisons of late-life cognitive aging. For up-to-date information, see https://g2aging.org.

## 7 Approach

**Overview:** This study develops and validates a **Sheaf-Theoretic Machine Learning (Sheaf-ML)** framework for multimodal integration and infection-aware cognitive risk prediction in Alzheimers Disease (AD). The approach unifies heterogeneous biomedical dataincluding serological infection markers, cardiovascular and metabolic parameters, behavioral and cognitive assessments, and wearable biosensor signalswithin a mathematically coherent **fibered multimodal data space**. Building on promising preliminary results from the **Harmonized LASI-DAD dataset**, we demonstrate that sheaf-based architectures can robustly model cross-domain dependencies, recover meaningful patient-level patterns despite missingness, and outperform conventional early-fusion baselines. At its core, Sheaf-ML represents multimodal data as sections of a sheaf defined over a topological domain of patients, where each modality constitutes a local chart. Learnable *restriction maps* encode inter-modality relationships, while a *gluing-consistency loss* enforces coherence among overlapping domains. Logic-regularized losses embed mechanistic priorssuch as the hypothesized causal link between infection burden and cognitive declineinto the learning objective, yielding biologically interpretable representations. The resulting integrated latent space enables computation of the **Infection Burden Index (IBI)**, a sheaf-derived, dynamic biomarker quantifying infection-related neurocognitive risk. The methodological trajectory spans three interlinked components: **1. Foundational Modeling:** Formulate and implement the Sheaf-ML architecture, defining patient-level fibers, modality domains, and topological interconnections. Validate the model on the Harmonized LASI-DAD dataset to benchmark sheaf-integrated embeddings against traditional machine learning models. **2. Predictive Validation of IBI:** Derive and evaluate the Infection Burden Index across multi-cohort datasets (ADNI, OASIS, NACC, ICMR, AMPAD). Assess reproducibility, robustness to missingness, interpretability, and alignment with infectioncognition pathways. **3. Clinical Translation and Adaptive Design:** Develop a pilot clinical study (NIMHANSLASI) to evaluate the clinical utility of IBI-guided interventions. Design adaptive feedback mechanisms linking infection monitoring, wearable biosensing, and cognitive assessment for real-time decision support. Together, these components establish a mathematically grounded and clinically interpretable framework for mechanism-informed dementia stratification. By coupling rigorous categorical topology with empirical validation and translational design, this approach lays the groundwork for a new generation of scalable, data-driven, and biologically meaningful models for Alzheimers Disease risk prediction and intervention.

### 7.1 Research Strategy

**1. Mathematical Foundation for Integration: 1.1 Sheaf Theory:** Biomedical data from EEG, wearables, and serology are inherently heterogeneous and temporally asynchronous. Conventional fusion methods often collapse these data into flat embeddings, erasing modality-specific structure and semantic context. Sheaf theory provides a principled mechanism for integrating such diverse signals by *gluing* locally defined observations into coherent global representations. Each modality is modeled as a local section over a domain of observation, while consistency maps ensure that overlaps are harmonized across modalities. This preserves the internal meaning of each data type and enables accurate, interpretable, patient-specific modeling. ^**11**^ **1.2 Topological Gluing:** Infection dynamics and neurodegeneration exhibit overlapping biological pathways, yet fragmented statistical models rarely capture these shared domains. Topological gluing uses set intersections to formally represent and connect data across these overlapping regionssuch as immune-inflammatory and neurophysiological domainsthereby creating a more biologically coherent model of cognitive decline. This structure allows infection-related and neurodegenerative signals to inform each other, improving model robustness and clinical interpretability.^**12**^ **1.3 Fibered Categories:** Patient trajectories evolve across time and modalities, creating longitudinal structures that conventional flattened models fail to capture. Fibered categories generalize sheaf-theoretic integration by treating each patients data as a structured *fiber* over a shared base category representing clinical progression. This abstraction maintains a global understanding of disease evolution while preserving within-patient variation and modality-specific dynamics.^**8, 9**^ Unlike traditional models that oversimplify patient heterogeneity, fibered categories provide a modular and scalable formalism that supports individualized disease trajectory modeling and explainable inference.^**13, 14**^ **1.4 Why These Mathematical Tools are Especially Suited:** Standard early-or late-fusion architectures often lose key inter-modal dependencies and cannot handle real-world data challenges such as noise, missingness, or temporal misalignment.^**15–19**^ In contrast, the categorical-topological framework is modular, hierarchical, and logically composable. Sheaf theory maintains local temporal and modality-specific structures, topological gluing aligns overlapping domains (e.g., immuneneural intersections), and fibered categories represent patient-specific trajectories as structured mappings. Together, these principles enable Sheaf-ML to model heterogeneous, asynchronous, and incomplete biomedical data in a mathematically consistent and biologically meaningful way, preserving semantic fidelity while ensuring robustness and generalizability. **2. Multimodal Data Anchoring Biological Reality: 2.1 EEG:** EEG captures high-temporal-resolution neural dynamics, serving as a sensitive marker of cognitive state fluctuations. Within the sheaf framework, EEG signals define local sections that reflect brain functional states, dynamically integrated with slower-varying physiological and serological data to detect subtle cognitive transitions in real time.^**1 2.2**^ **Wearables:** Wearable sensors provide continuous behavioral and physiological measures (e.g., heart rate variability, temperature, sleep architecture), offering a non-invasive window into systemic stress and inflammation. Despite their asynchrony and noise, sheaf-based fusion allows robust incorporation of these signals into the global data manifold, linking peripheral physiology to central neural changes.^**11**^ **2.3 Serology:** Serological assays provide episodic yet crucial insights into immune status and latent infection burden. Within Sheaf-ML, serological data anchor the biological basis of infection-modulated neurocognitive decline, linking HSV-1 or CMV seropositivity and antibody titers to changes in neural and behavioral metrics.^**20**^ **3. ML Integration for Patient-Level Prediction: 3.1 Feature Learning:** Unlike conventional black-box feature extraction, Sheaf-MLs structured representations are mathematically constrained by gluing consistency and fiber morphisms, producing interpretable latent features that encode biologically meaningful relationships across modalities.^**12**^ **3.2 Explainability:** A fibered-category representationwhere each modality forms a structured fiber projecting onto a shared patient-centered baseenables traceable and composable inference. This architecture allows clinicians to trace predictions to specific input modalities and physiological processes, enhancing clinical trust and explainability.^**1**^ **3.3 Patient-Level Prediction:** Sheaf-ML supports individualized risk modeling by maintaining the longitudinal coherence of each patients data fiber. The resulting **Infection Burden Index (IBI)** dynamically quantifies infection-related neurocognitive risk, allowing real-time tracking and personalized intervention strategies.^**2**^ **4. Domain Relevance and Translational Alignment: 4.1 Neuropharmacology:** Coupling biosensor and infection data with pharmacodynamic responses enables mechanistic insight into how infection burden modulates cognitive resilience or treatment efficacy.^**11**^ **4.2 Pharmacogenomics:** Incorporating polygenic risk scores and geneenvironment interactions allows stratification of individuals with differential susceptibility to infection-triggered neurodegeneration, enhancing precision in therapeutic targeting.^**20**^ **4.3 Bioethics & Clinical Trials:** Sheaf-MLs modular and categorical structure inherently supports ethical and transparent AI design by encoding constraints on consent, privacy, and fairness. These principles align with regulatory and clinical trial standards, ensuring responsible and equitable deployment.^**1**^

### 7.2 Sheaf-ML Framework: Categorical-Topological Modeling of Multimodal Data

We developed a sheaf-theoretic and categorical framework for integrating heterogeneous biomedical data modalities relevant to Alzheimer’s Disease (AD) progression modeling, specifically EEG signals, wearable sensor outputs, and serological biomarkers. Each modality is modeled as a local patch (open set) in a topological space, with overlaps encoding relationships and redundancies across data sources. A sheaf ℱ assigns to each patch the corresponding data (local section), and gluing conditions ensure global consistency. Furthermore, fibered categories abstract these assignments into a patient-level base category 𝒞, representing unified clinical interpretations.

### Visualization of the Sheaf-ML Framework

**First Row (Local Observations): (a) EEG Patch** *U*_1_: Local brainwave signal readings forming the EEG modality patch. **(b) Wearable Patch** *U*_2_: Biosensor measurements (heart rate, temperature) collected through wearables. **(c) Serology Patch** *U*_3_: Blood-based serological measures indicative of infection burden or immune status. **Second Row (Multimodal Integration): (d) Overlaps and Gluing:** Intersecting regions reflect shared or complementary physiological information across modalities, enabling topological gluing conditions. **(e) Sheaf Structure:** Sheaf ℱ assigns local sections (data) to each open set, ensuring compatibility under intersections and enabling global reconstruction. **(f) Fibered Category Representation:** Each modality-specific view (fiber) projects onto a base category 𝒞, representing a coherent, patient-centric clinical model. **Summary:** Local modality patches model heterogeneous sources of information. Overlaps ensure data consistency and alignment across modalities. Sheaf assignments enable structured multimodal fusion. Fibered category abstraction supports patient-level reasoning and machine learning integration.

### Mathematical Formulation

#### Statistical and ML Analysis

Machine learning will be performed in the sheaf neural network and fibered learner framework described in the mathematical formulation. Classical performance metrics (AUC, accuracy, F1-score) will be computed via cross-validation. Statistical comparison between MCI and controls will use t-tests, ANOVA, and chi-square tests. Multiple regression will model the impact of specific pathogen exposure and physiological shifts on cognitive decline.

#### Sheaf-ML Integration Feasibility

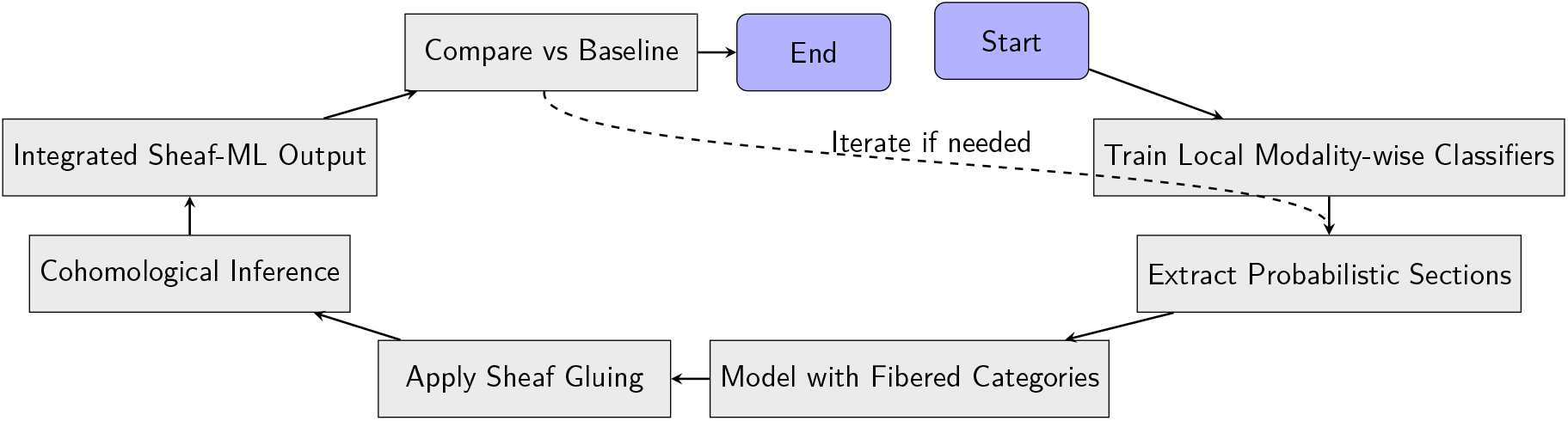

The process begins with training local classifiers independently on each modality (e.g., EEG, serology, wearables). These models generate probabilistic sections over the data space. The sections are structured into a fibered category, enabling mathematical organization of uncertainty and local-global correspondence. Through sheaf-ML gluing (e.g., Seifert–van Kampen), local predictions are integrated respecting topological and logical consistency. The resulting cohomological inference captures hidden interactions and multimodal interplay. The final sheaf-ML predictions are compared with baseline outputs (e.g., early fusion or RNNs), and the pipeline is refined iteratively.

### 7.3 Results: Multimodal Integration and Clinical Interpretability

**Overview** Understanding complex health outcomes such as cognitive decline and infection susceptibility requires integrating diverse sources of patient information. Individual health domainsdemographics, cognition, mental health, cardiovascular status, nutrition, and infection historyprovide complementary insights, but analyzing them independently often overlooks cross-domain interactions that drive clinically relevant outcomes. To address this challenge, we applied a sheaf-based framework to the Harmonized LASI-DAD dataset, enabling systematic integration of these heterogeneous modalities into a unified, patient-centered representation.

Our approach is motivated by three key considerations: 1. **Capturing Multimodal Dependencies:** Traditional models treat features as independent or require manual feature engineering to account for inter-domain interactions. By leveraging sheaf-theoretic structures, we can naturally encode dependencies between modalities, allowing data from one domain (e.g., infection biomarkers) to inform and contextualize signals from others (e.g., cardiovascular or cognitive measures). 2. **Maintaining Topological Consistency:** Patient data are inherently structured; each individual represents a local neighborhood of correlated features. Sheaf-based integration respects this local-to-global structure, ensuring that modality-specific embeddings are coherently aligned when projected into a shared patient-level space. This preserves the intrinsic organization of the data while supporting systematic cross-domain reasoning. 3. **Enabling Clinically Interpretable Machine Learning:** By creating a topologically integrated dataset, we provide a foundation for downstream predictive modeling (Sheaf-ML) that is both accurate and interpretable. The model can identify global cohort-level drivers, as well as patient-specific risk patterns, supporting actionable insights for early detection and personalized intervention.

In this study, *N* = 6168 participants with complete multimodal data were included. Six biologically and behaviorally relevant domainsdemographics, cognition, mental health, cardiovascular function, nutrition, and infectionwere mapped into a sheaf-based framework, producing a 111-dimensional integrated representation for each patient. This representation captures both intra-modality information (e.g., detailed cognitive test scores) and cross-modality dependencies (e.g., how infection burden affects cardiovascular and cognitive outcomes).

Through this integration, our framework achieves two goals simultaneously: - *System-level understanding* : revealing biologically meaningful coordination among multiple health domains, particularly the central role of infection burden in linking physiological and cognitive processes. - *Individual-level insight*: enabling personalized clinical interpretation through patient fibers that highlight neighborhood-level similarity and heterogeneity in multimodal health profiles. Overall, this approach provides a mathematically principled, clinically relevant framework for analyzing complex health datasets, bridging the gap between sophisticated topological modeling and actionable medical insights.

#### I.

**Multimodal Data Integration**

We analyzed six key domains: **Demographics** (age, sex, education; 5 features), **Cognition** (memory and cognitive tests; 64 features), **Mental Health** (mood and stress indicators; 8 features), **Cardiovascular** (blood pressure, heart rate, etc.; 24 features), **Nutrition** (diet-related measures; 6 features), and **Infection** (history and biomarkers; 4 features). A total of *N* = 6168 participants were included.

Using sheaf-based integration, we aligned these domains into a single patient-level representation, capturing how features from one domain influence others. For instance, integrating infection data revealed clinically meaningful links between infection, cardiovascular health, nutrition, and cognitive performance (Figure 6).

Figures 4 and 5 quantify each domain’s contribution to infection burden at both population and patient levels, highlighting coordinated influences of infection, cardiovascular, and cognitive features.

**Figure 1.**
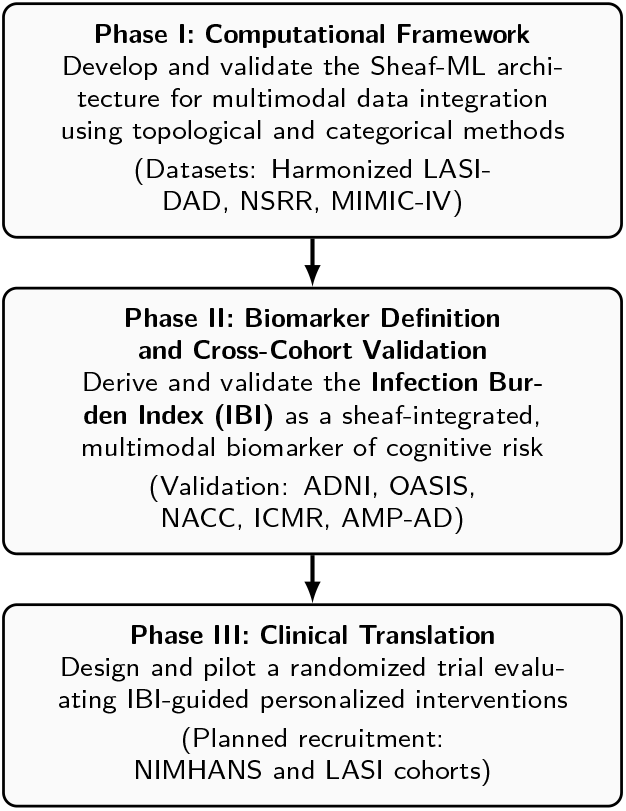
Schematic roadmap illustrating the phased development of the Sheaf-ML and Infection Burden Index (IBI) framework, progressing from computational modeling and validation toward clinical translation.

**Figure 2.**
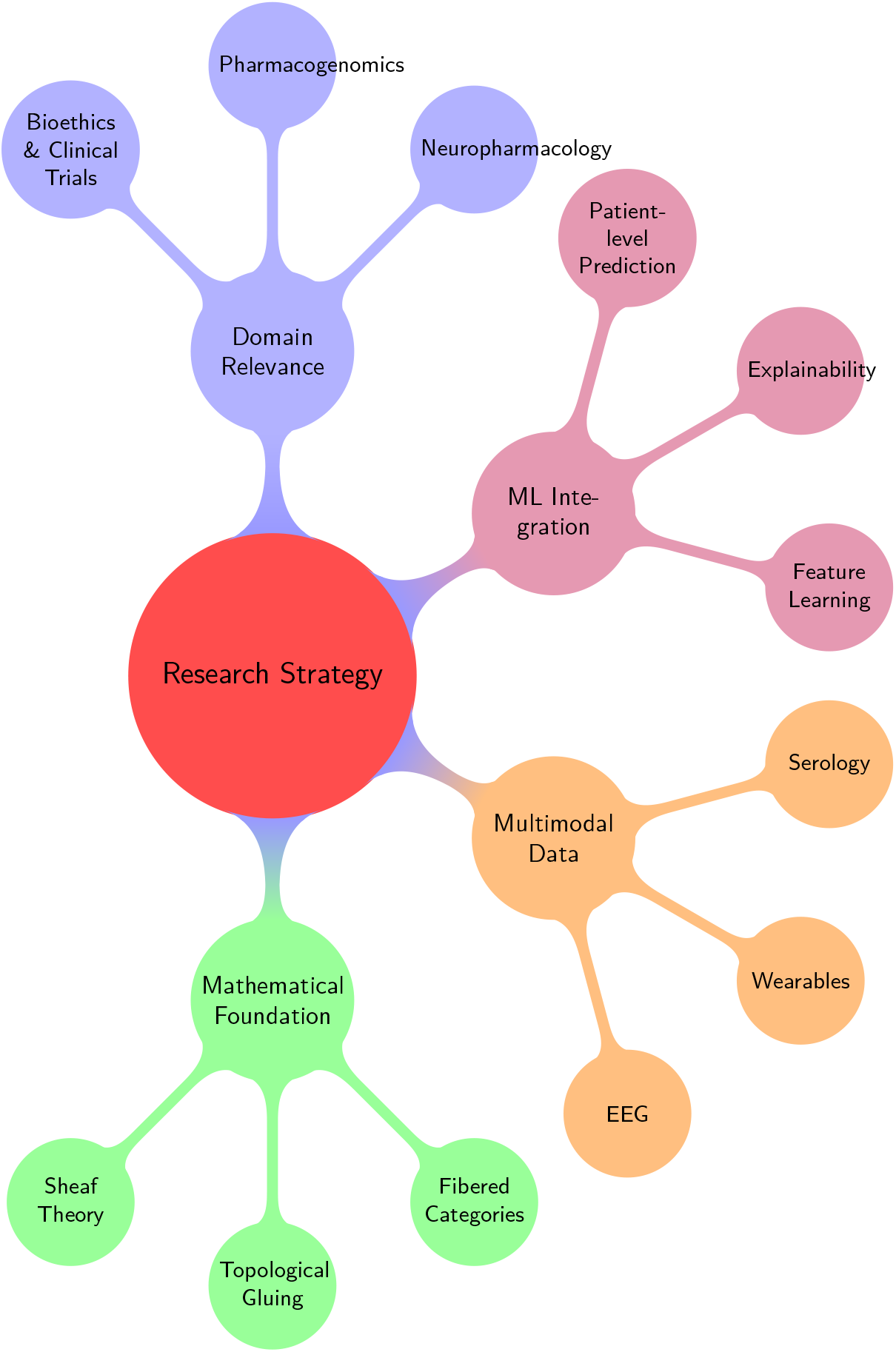
Conceptual structure of the research strategy, integrating mathematical foundations, multimodal data, ML modeling, and domain alignment.

**Figure 3.**
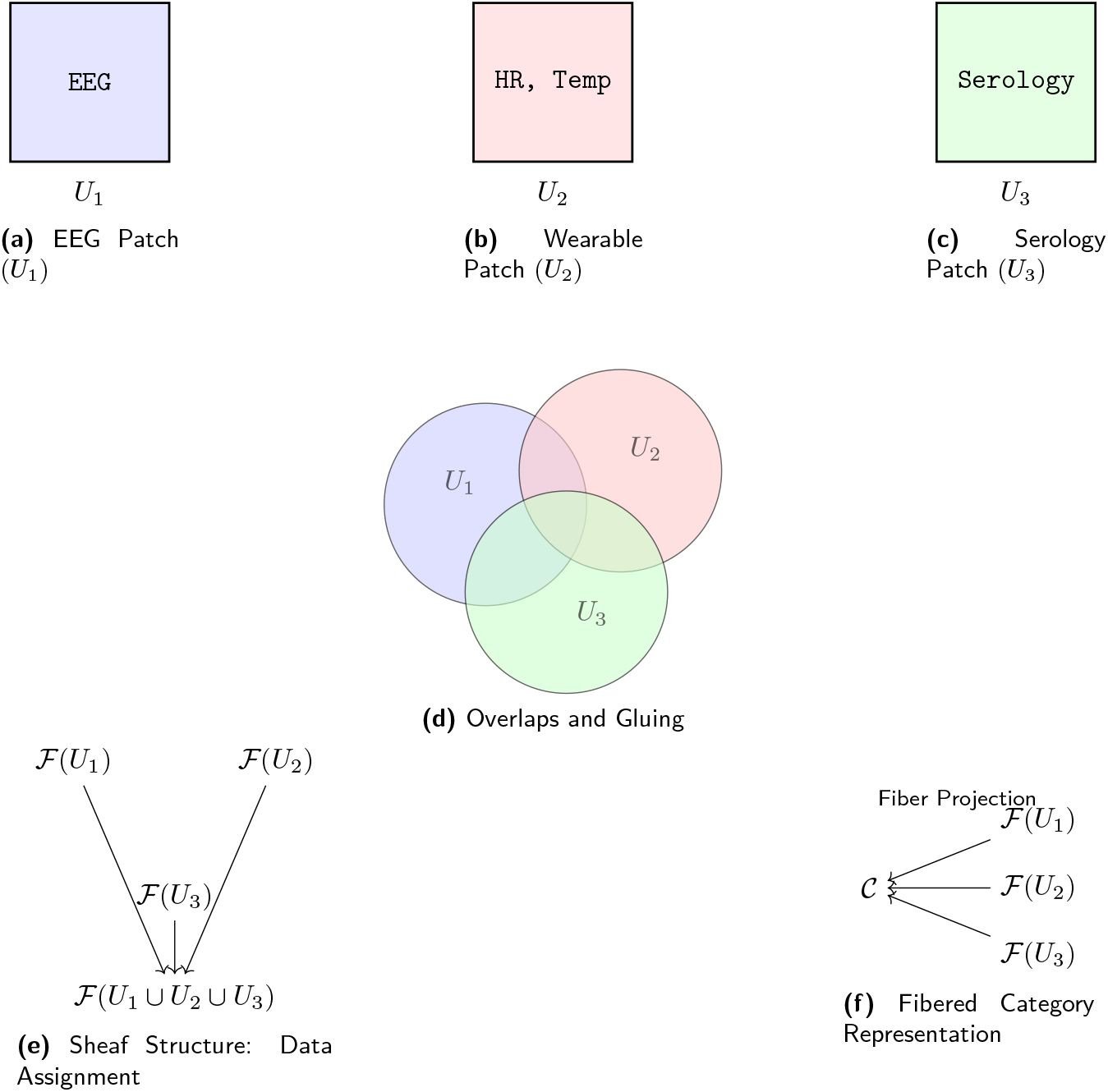
Categorical and Topological Integration of Multimodal Data:EEG, wearable, and serological data are modeled as open sets *U*_*i*_ with local sections ℱ (*U*_*i*_) assigned via a sheaf. Overlaps encode topological gluing, while the fibered category projects local data to a global patient-level integration 𝒞.

**Figure 4.**
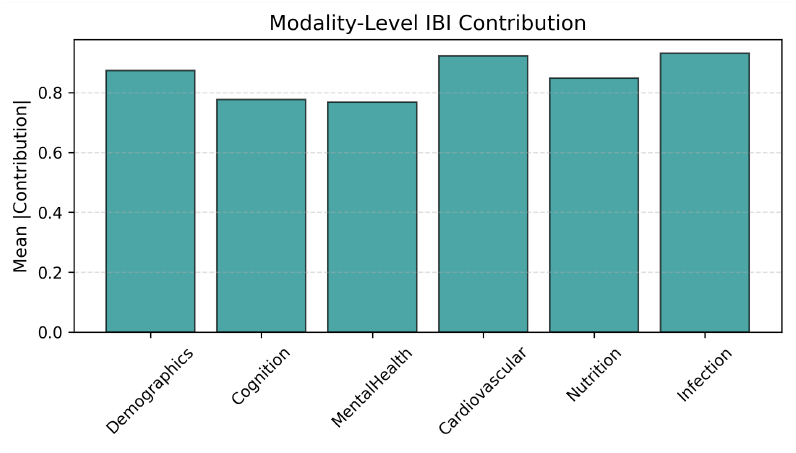
Modality-Level IBI Contribution: relative influence of each domain on infection burden.

**Figure 5.**
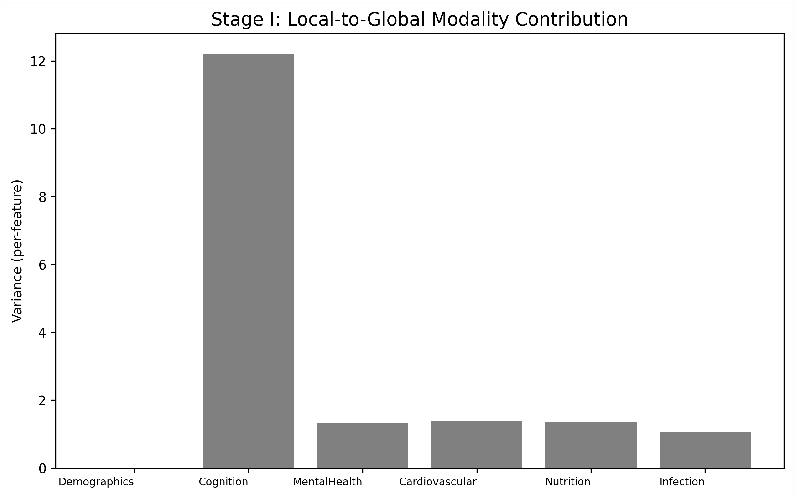
Local-Global IBI Contribution: domain-level signals propagated into global patient embeddings.

**Technical Explanation: Categorical and Topological Integration**

We define six biologically grounded modalities ℳ corresponding to feature subsets of the 111-dimensional space. Each modality *M*_*i*_ ∈ ℳ is assigned a presheaf ℱ i mapping patient or local neighborhoods to observed data, supporting restriction maps *ρUV* that encode inter-domain dependencies. Sheaf consistency is enforced via a gluing condition, penalizing discrepancies in overlapping domains.

Globally, each patient defines a fiber ℰ_*b*_ populated by modality-specific sheaves. The cohort-level assembly forms an object in the topos of sheaves Sh(X), producing an integrated dataset. Comparison of baseline and IBIinformed networks (Fig. 6) shows infection burden introduces edges *linking Infection, Cardiovascular, Cognition, and Nutrition*, revealing biologically meaningful coordination.

**Figure 6.**
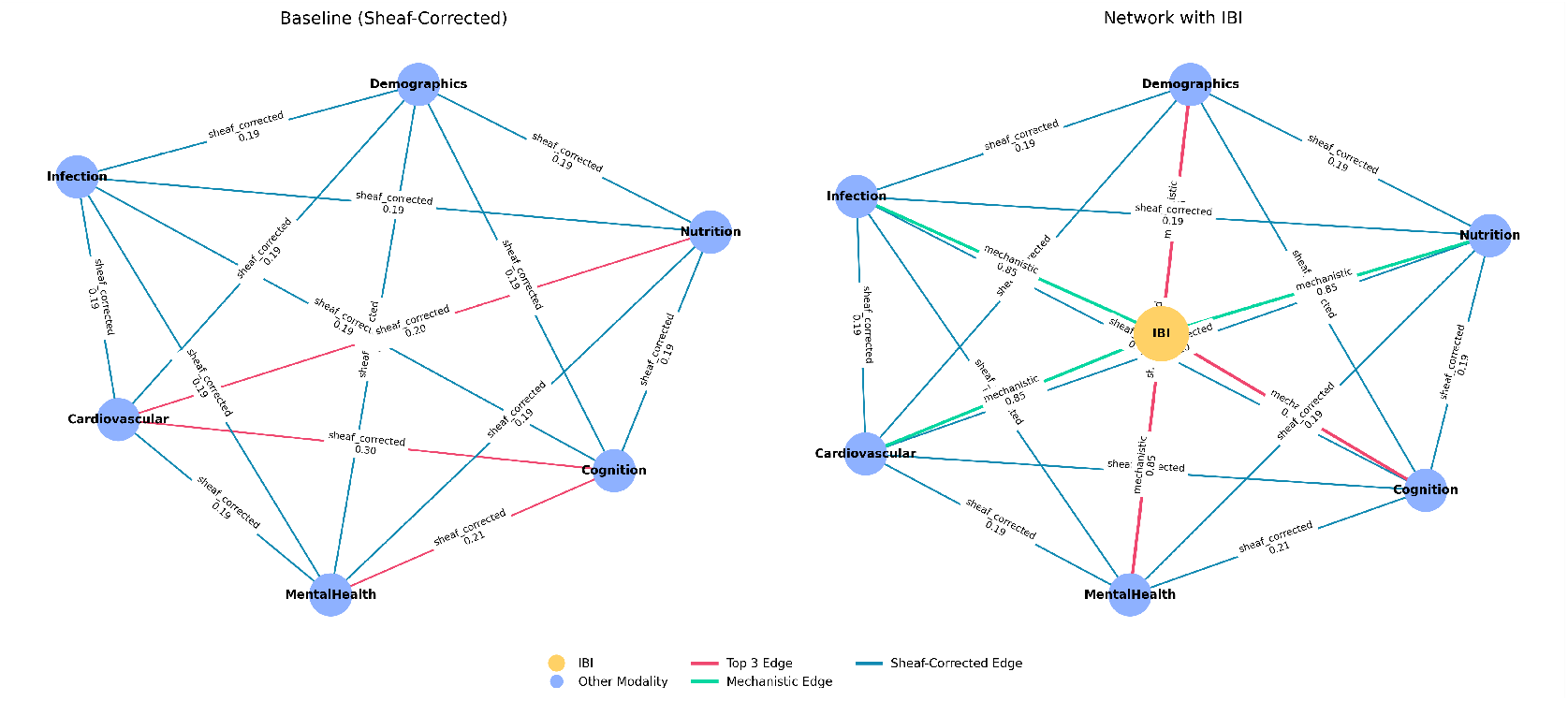
Baseline vs. IBI Sheaf-Integrated Multimodal Graph: infection burden introduces new links between domains, reflecting clinically meaningful coordination. Details are provided in the SheafML+IBI Clinical Report.

Figures 4 and 5 quantify modality-level and localglobal contributions, demonstrating that sheaf-based integration aligns feature spaces while preserving clinically interpretable cross-domain interactions. This integrated representation enables downstream machine learning to leverage system-level patterns for prediction of cognitive vulnerability and infection-related risk

#### II.

**Learning from Integrated Data: Sheaf-ML**

Using the sheaf-integrated patient representation as input, **Sheaf-ML** captures both within-domain and crossdomain interactions.

Global feature contributions identify the most influential predictors cohort-wide, while patient-level contributions highlight individualized patterns of cross-domain influence. Patient fiber contexts visualize neighborhood-level similarity, facilitating identification of stable versus heterogeneous risk patterns.

**Technical Explanation: Sheaf-ML**

We construct an **Integrated Sheaf Dataset** 𝒟 _int_ representing the sheaf-integrated embedding space X ∈ ℝ ^6168×111^, encoding both intra-and inter-modality fusion.

**Sheaf Neural Network:** The model applies a two-layer sheaf neural network, propagating embeddings across modality overlaps:

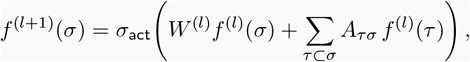

where *f* ^(*l*)^ (*σ*) is the embedding of domain *σ* at layer *l*.

**Consistency and Logic Regularization:** A gluing-aware term enforces *f* ^(*l*)^ (*σ*)|_τ_ ≈ *f* ^(*l*)^ (τ), and a logic-regularized loss incorporates domain priors such as higher infection burden implies elevated cognitive risk. The total loss is:

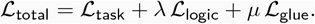

#### III.

**Clinical Interpretation: Infection Burden Index (IBI)**

Each patients cumulative infection-related risk was quantified using the IBI. Patients exceeding the 80^th^ percentile (*N* = 1234) were flagged as early-warning cases. High IBI scores were primarily driven by infection biomarkers, followed by cardiovascular and cognitive features (Fig. 10).

**Structural and Clinical Interpretability:** Global feature contributions (Fig. 7) highlight cohort-level drivers, patient-level contributions (Fig. 8) reveal individualized cross-domain influences, and patient fiber contexts (Fig. 9) illustrate neighborhood-level embeddings. Homogeneous neighborhoods indicate stable predictions, while heterogeneous fibers suggest atypical presentations requiring closer evaluation. These results demonstrate that Sheaf-ML not only achieves high predictive performance but also provides interpretable insights at both population and individual levels, supporting early detection and personalized stratification while preserving multimodal topological structure.

**Figure 7.**
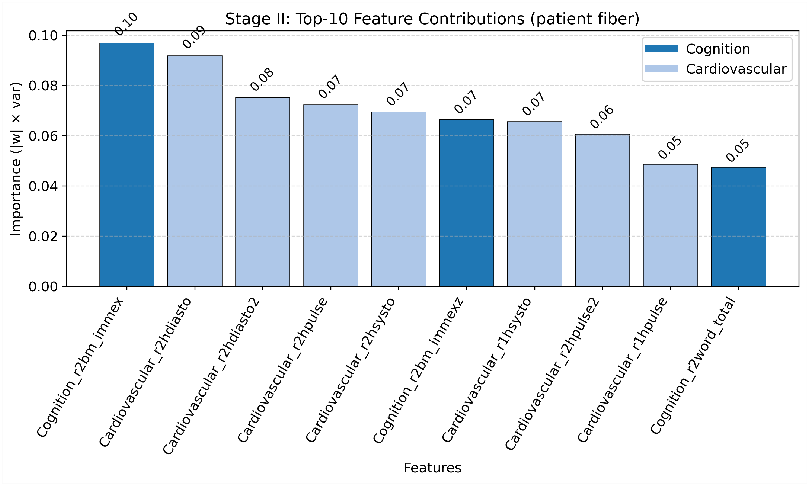
Global Feature Contributions: top predictors across the cohort.

**Figure 8.**
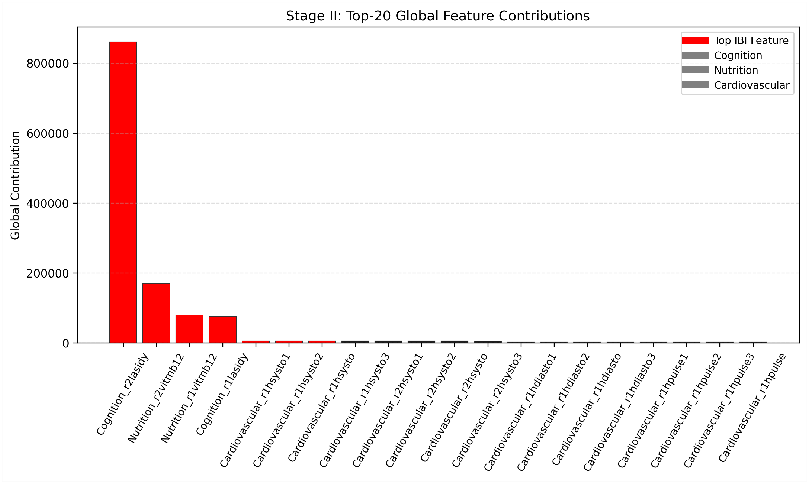
Patient Feature Contributions: individualized feature influence on outcomes.

**Figure 9.**
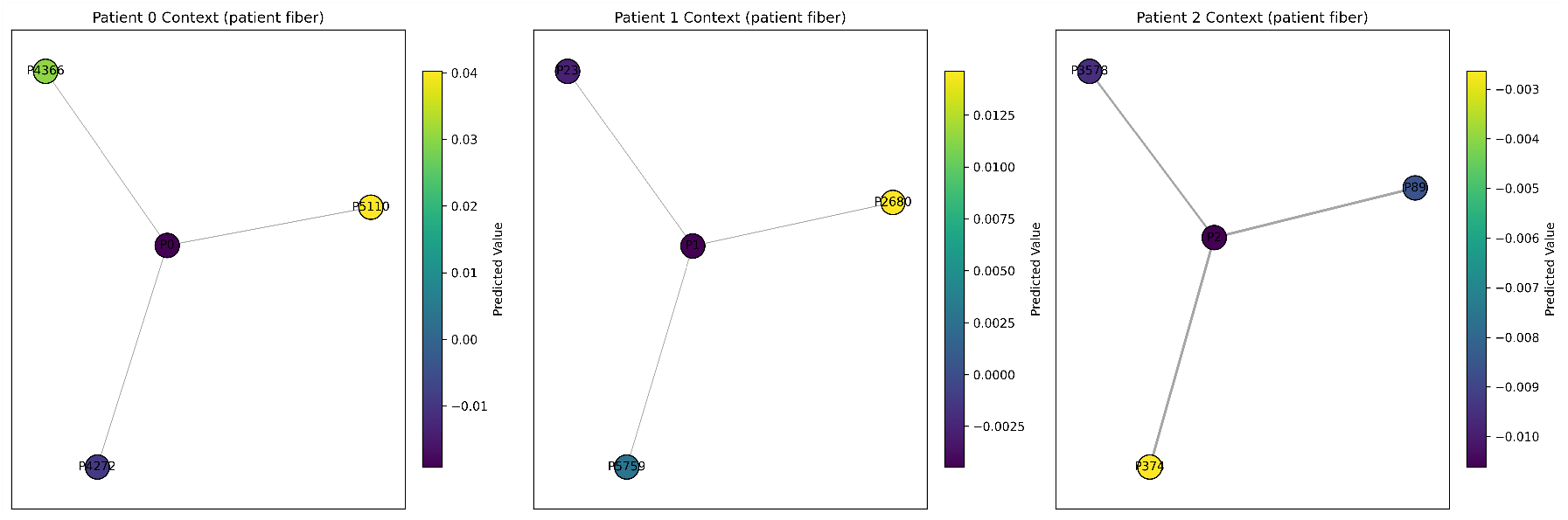
Patient Fiber Contexts: neighborhood graphs of patients with similar multimodal profiles. Node color indicates predicted health burden. Details are provided in the SheafML+IBI Clinical Report.

#### Summary

Our framework unifies multiple health domains into an interpretable patient representation, uncovers clinically meaningful interactions, and enables early detection of infection-related risk. By linking global patterns with patientspecific insights, it facilitates both population-level analysis and personalized clinical decision-making.

### 7.4 Potential Pitfalls, Challenges, and Alternative Strategies

Despite leveraging rigorous mathematical modeling and preliminary evidence for multimodal integration, several practical and methodological challenges are anticipated. Here, we outline key potential pitfalls and corresponding mitigation strategies to ensure model robustness, interpretability, and translational feasibility. **1. Sparse or incomplete multimodal data:** Real-world datasets often exhibit missingness and irregular sampling across serological, physiological, and neurocognitive domains. The sheaf-based framework inherently accommodates partial observability by enforcing local consistency conditions and gluing across incomplete data domains. To further ensure resilience, we will perform simulated missingness experiments and evaluate model performance under varying levels of data sparsity. **2. Cohort variability and population heterogeneity:** Differences in ancestry, socioeconomic background, or data acquisition protocols can introduce cohort-level biases. The fibered-category structure of our framework enables patient-specific embeddings that are robust to population heterogeneity, ensuring that learned relationships remain interpretable and generalizable across diverse subgroups. **3. Data heterogeneity and measurement variability:** Integration of biosensing data from diverse sourcessuch as wearable devices, EEG, or lab assaysmay introduce inconsistencies. Sheaf-based harmonization, combined with gluing-aware regularization and fibered modeling, mitigates these effects by accounting for environmental, contextual, and device-related variability, thereby preserving the fidelity of cross-domain interactions. **4. Longitudinal attrition and compliance risk:** Extended monitoring studies can face participant dropout or irregular engagement, particularly with wearable or smartphone-based assessments. To address this, the study incorporates structured participant engagement, adherence incentives, and robust temporal interpolation using categorical patching techniques that maintain continuity in multimodal embeddings. **5. Interpretability versus model complexity:** Highdimensional, multimodal integration can increase model complexity, potentially reducing interpretability. Our approach mitigates this risk by explicitly constructing patient fibers, modality-level contributions, and global-to-local interaction maps, allowing transparent, clinically meaningful reasoning even in complex datasets.

#### Summary

Collectively, these strategies strengthen the translational pipeline by ensuring data integrity, generalizable predictive performance, and reliable patient-level interpretation. By combining mathematical rigor, topological consistency, and clinical grounding, the framework provides a robust foundation for delivering a biologically interpretable AI system capable of predicting Alzheimers disease progression and infection-related risk across diverse populations.

## 8 Discussion

This work introduces a mathematically grounded framework**Sheaf-ML**for integrating heterogeneous biomedical data to model infection-related cognitive and systemic risk. By combining tools from algebraic topology and category theory with multimodal machine learning, we demonstrate that structurally informed integration enables coherent synthesis of serological, physiological, cognitive, and mental health data streams, which are often asynchronous, noisy, and incomplete. The resulting **Infection Burden Index (IBI)** quantifies cumulative infection-related risk at the patient level and provides a clinically interpretable biomarker for early, personalized intervention (Figure 10).

**Figure 10.**
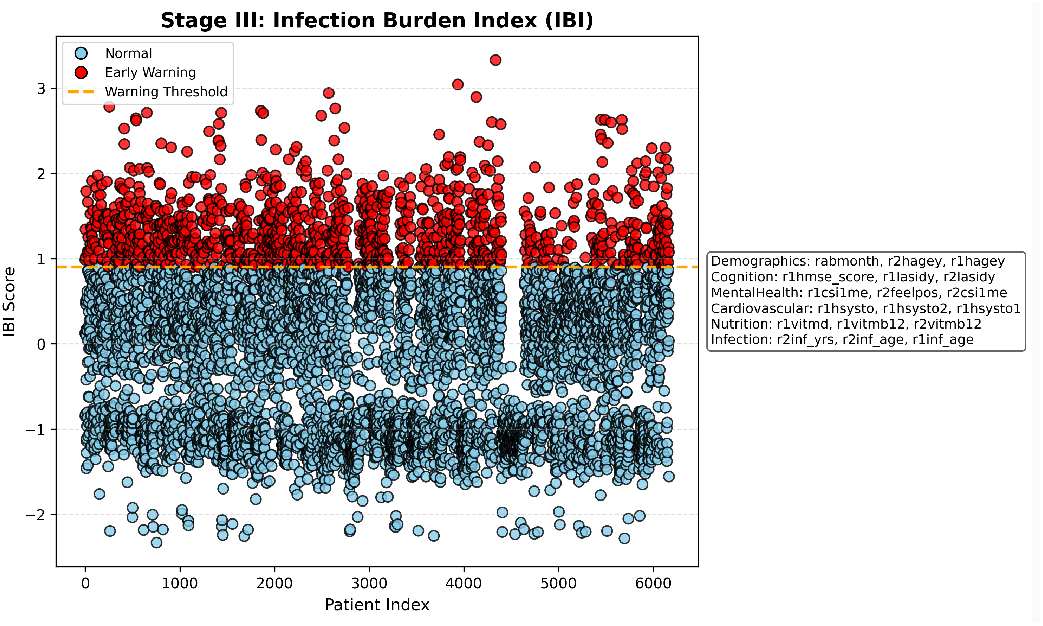
Patient-level IBI scores; top 20% flagged as early-warning cases. Details are provided in the SheafML+IBI Clinical Report.

Our results indicate that the sheaf-based approach resolves several limitations of conventional multimodal fusion. Traditional early-or late-fusion pipelines treat heterogeneous biomedical signals as flat vectors, erasing domain-specific relationships and reducing interpretability. In contrast, Sheaf-ML preserves the contextual geometry of each modality through local sections and restriction maps, while aligning overlapping domains via topological gluing. This framework captures meaningful cross-domain interactions, such as coordinated influences of infection, cardiovascular, and cognitive features, which were revealed in the IBI-driven network analysis (Figures 4 – 6). The fibered-category formulation further enables patient-specific modeling, linking global cohort patterns with individualized trajectories (Figures 7 – 8).

By embedding infection biomarkers directly into the model structure, Sheaf-ML operationalizes mechanistic hypotheses relating chronic infection to neurocognitive decline. The IBI does not function merely as a statistical risk score; it represents a topologically grounded summary of cross-domain interactions, highlighting patients with high infection burden and associated cardiovascular and cognitive vulnerability (Figure 10). Patient fiber context analysis additionally identifies clusters of patients with similar multimodal profiles, supporting personalized risk stratification and early-warning detection (Figure 9).

The translational potential is substantial. In clinical settings, Sheaf-ML could support adaptive monitoring platforms that integrate serological data, physiological measures, and cognitive assessments. By providing both population-level insights and patient-specific interpretations, it enables early identification of high-risk trajectories and informs interventions before irreversible neurodegeneration occurs. Moreover, the models explainable and modular design supports ethical deployment and regulatory compliance, addressing a key limitation of existing AI-based clinical tools.

Limitations include the reliance on cross-sectional, harmonized datasets; prospective validation is required to confirm predictive performance across diverse populations. Although Sheaf-ML is robust to missingness and noise, measurement heterogeneity remains a practical challenge, particularly in low-resource settings. Additionally, the current focus on infection-driven pathways should be extended in future work to incorporate complementary mechanisms such as vascular, metabolic, and genetic risk factors.

## 9 Future Steps

Building on the multimodal integration and patient-level interpretability demonstrated in this work, future directions will focus on clinical translation and methodological expansion: **Multimodal Expansion:** Extend Sheaf-ML to incorporate high-dimensional omics (transcriptomics, proteomics, metabolomics) and neuroimaging modalities. This will enhance mechanistic interpretability and enable finer-grained mapping of infection, immune dysregulation, and neurodegenerative pathways, leveraging insights from domain-level contributions (Figures 4 and 5). **Personalized Therapeutics:** As a future extension, we propose integrating pharmacogenomic and multi-ancestry polygenic risk scores (MAP-PRS)^**21**^ into the IBI framework, alongside longitudinal treatment-response data. This approach enables individualized therapeutic recommendations that account for each patients multimodal profile, infection-related cognitive risk, and genetic predisposition. By incorporating genetic risk as an additional modality, Sheaf-ML can model intra- and cross-modality interactions, supporting precision interventions and equitable, ancestry-aware treatment strategies (Figures 8 and 9). **Clinical Translation:** Deploy IBI-guided pilot clinical studies using cohorts such as **NIMHANS** and **LASI-DAD**. These studies will evaluate the feasibility, clinical impact, and decision-support potential of continuous infection monitoring and early intervention in cognitive decline and dementia prevention (Figure 10).

These initiatives aim to transition Sheaf-ML from a computational prototype into a clinically deployable platform for infection-informed cognitive risk assessment and early-warning detection.

## 10 Conclusion

The **Sheaf-ML** framework and the derived **Infection Burden Index (IBI)** provide a conceptual and methodological advance in modeling cross-domain interactions underlying infection-related cognitive risk. By uniting rigorous topological and categorical mathematics with multimodal patient data, our approach enables interpretable, patient-specific, and population-informed insights into infectioncognition dynamics.

Preliminary results show that Sheaf-ML captures latent cross-domain interactions and identifies high-risk patients even with noisy or incomplete data, while patient fiber contexts allow stratification based on similar multimodal profiles (Figures 9, 7, 8). Future work incorporating high-dimensional omics, pharmacogenomic data, and IBI-guided clinical trials will strengthen translational applicability. If validated prospectively, this paradigm could shift dementia care from reactive diagnosis toward proactive, personalized preventionbridging theoretical rigor with clinical impact.

## Data Availability

Harmonized LASI-DAD programming codes and Codebook, Version B.1

https://g2aging.org

https://doi.org/10.25553/h5wx-ay45

## Acknowledgements

We are grateful to Dr. Eyal Y. Kimchi (Department of Neurology, Northwestern University Feinberg School of Medicine, USA), Dr. Strajit Ghosh (Massachusetts General Hospital and Harvard Medical School, USA), Dr. Fernando Gómez-Baquero (Jacobs TechnionCornell Institute, Cornell Tech; NSF Upstate New York Energy Storage Engine, USA), Dr. Kamana Porwal (Department of Mathematics, IIT Delhi, India), and Dr. Mustafa Hajij (MSDSAI Program, University of San Francisco, California, USA) for their valuable guidance, support, and insightful suggestions, which greatly contributed to this work.

## Author Contributions

-Dr. Lokendra S. Thakur conceived the idea, designed the study, developed the novel Sheaf-ML method and analysis, developed pipeline, all sections writing. - Dr. Gurpreet Bharj performed the clinical analysis. - Duc Thanh Nguyen checked citations and contributed in refining content. - Manish Saroya contributed in writing significance and innovation sections. - Bushra Malik contributed to data sources and availability section. - All authors reviewed and approved the final version.

## Ethics Approval and Consent to Participate

Not applicable.

## Data Availability

We accessed raw data from the publicly available repository: (https://g2aging.org) Code supporting this study are available on reasonable request.

## Disclaimer

Preprints are preliminary reports that have not been peer reviewed. They should not be regarded as conclusive, guide clinical practice, or be reported in news media as established information.

## SheafML + IBI Clinical Report

### Stage I: Local Embeddings

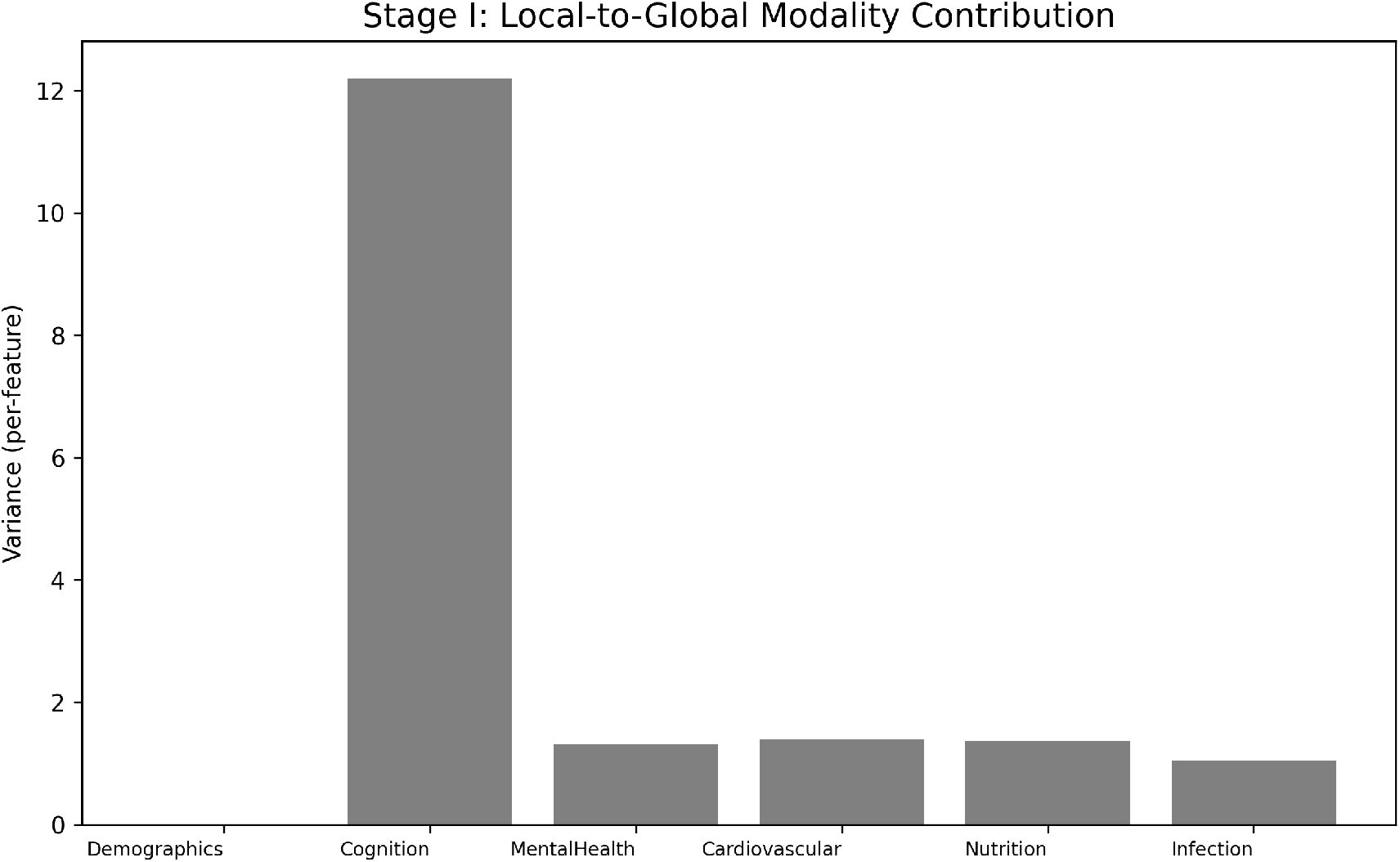

Demographics local features: pn, rabyear, rabmonth, r1hagey, r2hagey

Cognition local features: r1lasidy, r2lasidy, r1hmse_score, r2hmse_score, r1lasi_score, r2lasi_score, r1word1, r2word1, r1word2, r2word2, r1word3, r2word3, r1word_total, r2word_total, r1word_d, r2word_d, r1bm_imm, r2bm_imm, r1bm_imm_d, r2bm_imm_d, r1bm_immex, r2bm_immex, r1bm_recl, r2bm_recl, r1bm_recl_d, r2bm_recl_d, r1bm_reclex, r2bm_reclex, r1lmb_immex, r2lmb_immex, r1lmb_immg, r2lmb_immg, r1lmb_imm, r2lmb_imm, r1lmb_reclex, r2lmb_reclex, r1lmb_reclg, r2lmb_reclg, r1lmb_recl, r2lmb_recl, r1log_reco, r2log_reco, r1verbal, r2verbal, r1verbal_inc, r2verbal_inc, r1rv_score, r2rv_score, r1hmse_scorz, r2hmse_scorz, r1word_totaz, r2word_totaz, r1word_dz, r2word_dz, r1log_recoz, r2log_recoz, r1bm_immexz, r2bm_immexz, r1bm_reclexz, r2bm_reclexz, r1verbalz, r2verbalz, r1rv_scorez, r2rv_scorez

MentalHealth local features: r1csid_scorz_d, r2csid_scorz, r1feelpos, r2feelpos, r1feelneg, r2feelneg, r1csi1me, r2csi1me

Cardiovascular local features: r1hsysto1, r2hsysto1, r1hsysto2, r2hsysto2, r1hsysto3, r2hsysto3, r1hsysto, r2hsysto, r1hdiasto1, r2hdiasto1, r1hdiasto2, r2hdiasto2, r1hdiasto3, r2hdiasto3, r1hdiasto, r2hdiasto, r1hpulse1, r2hpulse1, r1hpulse2, r2hpulse2, r1hpulse3, r2hpulse3, r1hpulse, r2hpulse

Nutrition local features: r1vitmb12, r2vitmb12, r1folacid, r2folacid, r1vitmd, r2vitmd

Infection local features: r1inf_age, r2inf_age, r1inf_yrs, r2inf_yrs

### Stage II: Fused Features & Predictions

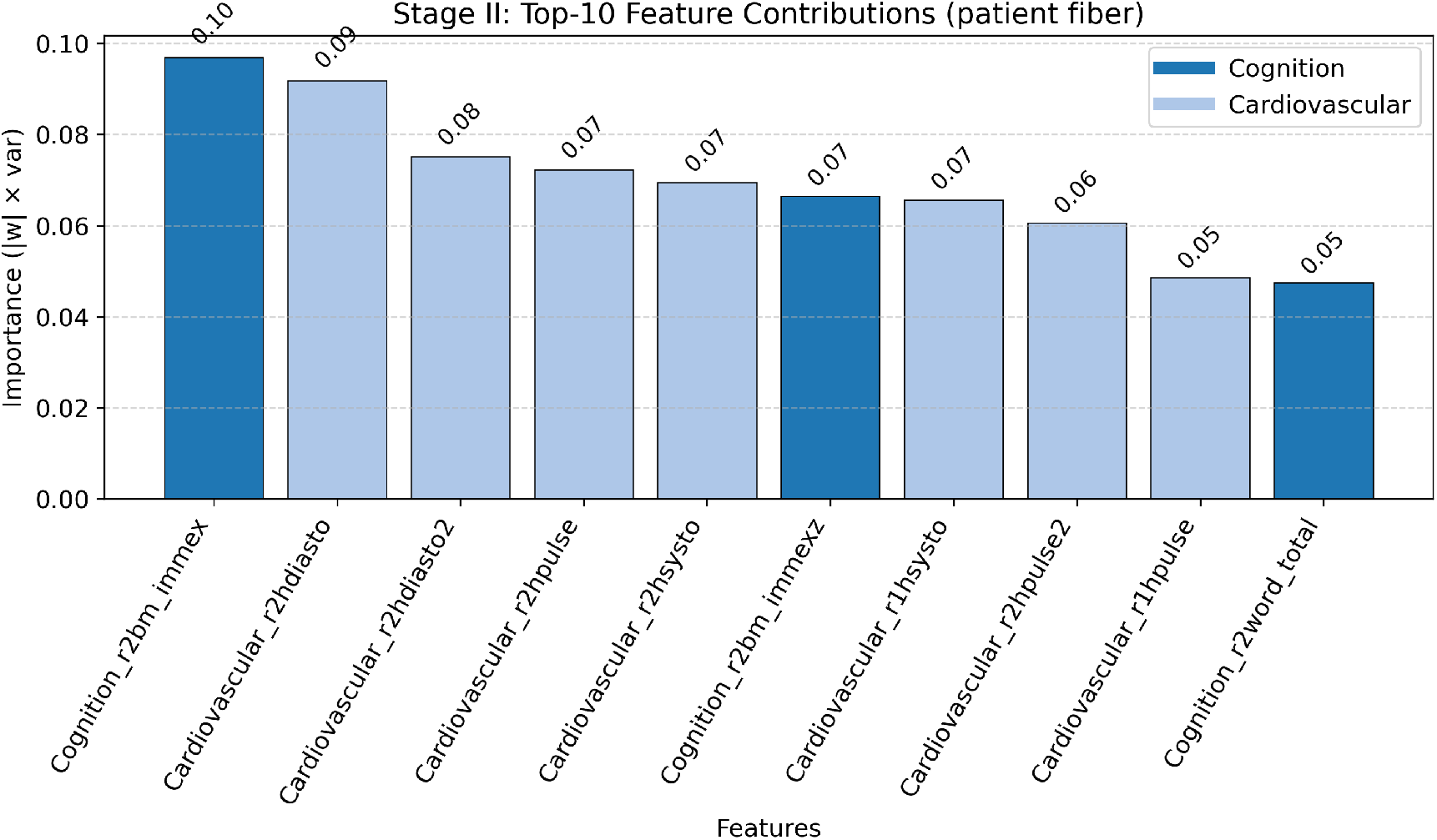

Demographics features: Demographics_pn, Demographics_rabyear, Demographics_rabmonth, Demographics_r1hagey, Demographics_r2hagey

Cognition features: Cognition_r1lasidy, Cognition_r2lasidy, Cognition_r1hmse_score, Cognition_r2hmse_score, Cognition_r1lasi_score, Cognition_r2lasi_score, Cognition_r1word1, Cognition_r2word1, Cognition_r1word2, Cognition_r2word2, Cognition_r1word3, Cognition_r2word3, Cognition_r1word_total, Cognition_r2word_total, Cognition_r1word_d, Cognition_r2word_d, Cognition_r1bm_imm, Cognition_r2bm_imm, Cognition_r1bm_imm_d, Cognition_r2bm_imm_d, Cognition_r1bm_immex, Cognition_r2bm_immex, Cognition_r1bm_recl, Cognition_r2bm_recl, Cognition_r1bm_recl_d, Cognition_r2bm_recl_d, Cognition_r1bm_reclex, Cognition_r2bm_reclex, Cognition_r1lmb_immex, Cognition_r2lmb_immex, Cognition_r1lmb_immg, Cognition_r2lmb_immg, Cognition_r1lmb_imm, Cognition_r2lmb_imm, Cognition_r1lmb_reclex, Cognition_r2lmb_reclex, Cognition_r1lmb_reclg, Cognition_r2lmb_reclg, Cognition_r1lmb_recl, Cognition_r2lmb_recl, Cognition_r1log_reco, Cognition_r2log_reco, Cognition_r1verbal, Cognition_r2verbal, Cognition_r1verbal_inc, Cognition_r2verbal_inc, Cognition_r1rv_score, Cognition_r2rv_score, Cognition_r1hmse_scorz, Cognition_r2hmse_scorz, Cognition_r1word_totaz, Cognition_r2word_totaz, Cognition_r1word_dz, Cognition_r2word_dz, Cognition_r1log_recoz, Cognition_r2log_recoz, Cognition_r1bm_immexz, Cognition_r2bm_immexz, Cognition_r1bm_reclexz, Cognition_r2bm_reclexz, Cognition_r1verbalz, Cognition_r2verbalz, Cognition_r1rv_scorez, Cognition_r2rv_scorez

MentalHealth features: MentalHealth_r1csid_scorz_d, MentalHealth_r2csid_scorz, MentalHealth_r1feelpos, MentalHealth_r2feelpos, MentalHealth_r1feelneg, MentalHealth_r2feelneg, MentalHealth_r1csi1me, MentalHealth_r2csi1me

Cardiovascular features: Cardiovascular_r1hsysto1, Cardiovascular_r2hsysto1, Cardiovascular_r1hsysto2, Cardiovascular_r2hsysto2, Cardiovascular_r1hsysto3, Cardiovascular_r2hsysto3, Cardiovascular_r1hsysto, Cardiovascular_r2hsysto, Cardiovascular_r1hdiasto1, Cardiovascular_r2hdiasto1, Cardiovascular_r1hdiasto2, Cardiovascular_r2hdiasto2, Cardiovascular_r1hdiasto3, Cardiovascular_r2hdiasto3, Cardiovascular_r1hdiasto, Cardiovascular_r2hdiasto, Cardiovascular_r1hpulse1, Cardiovascular_r2hpulse1, Cardiovascular_r1hpulse2, Cardiovascular_r2hpulse2, Cardiovascular_r1hpulse3, Cardiovascular_r2hpulse3, Cardiovascular_r1hpulse, Cardiovascular_r2hpulse

Nutrition features: Nutrition_r1vitmb12, Nutrition_r2vitmb12, Nutrition_r1folacid, Nutrition_r2folacid, Nutrition_r1vitmd, Nutrition_r2vitmd

Infection features: Infection_r1inf_age, Infection_r2inf_age, Infection_r1inf_yrs, Infection_r2inf_yrs

### Stage III: IBI Scores & Early-Warning Flags

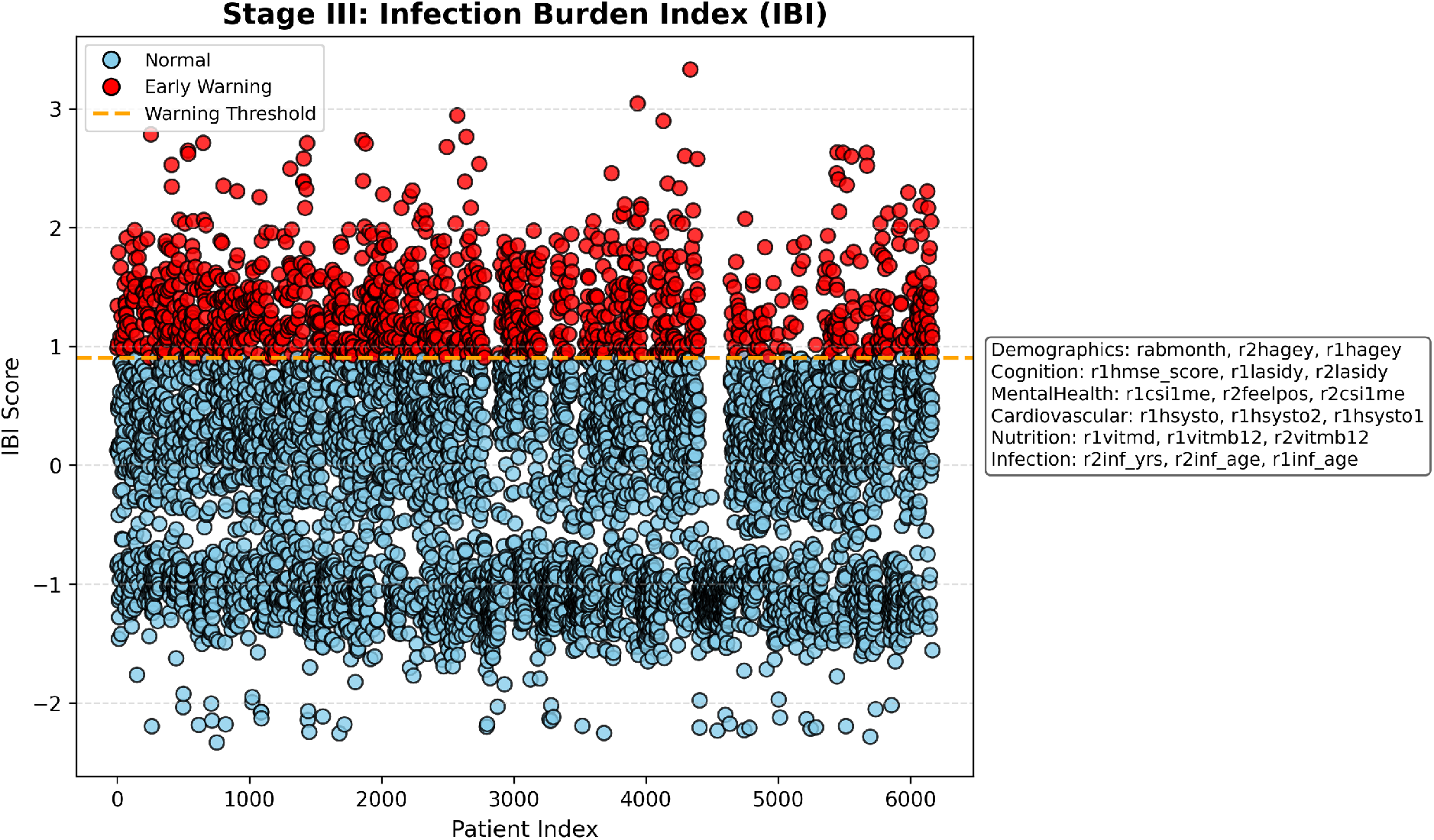

#### Early Warning Patients (IBI)

Number flagged: 1234 / 6168

